# Convergence for Inactivation of TGFβ Signaling Is a Common Feature of Advanced Pancreatic Cancer

**DOI:** 10.1101/2024.01.30.24301554

**Authors:** Jungeui Hong, Zachary Kohutek, Haochen Zhang, Elias-Ramzey Karnoub, Rajya Kappagantula, Laura D. Wood, Christine Iacobuzio-Donahue

## Abstract

We performed WES of 250 unique tumor tissues from 30 multiregion sampled pancreatic cancer research autopsies from patients diagnosed with advanced stage disease. We find that most genetic alterations in PDAC occur in a subclonal manner, and some genes occurred in a subclonal manner exclusively. Convergent evolution within the TGFβ pathway was also identified as a common feature of advanced stage disease, with *SMAD4* inactivation more common among metastatic PDACs compared to inactivation of TGFβ surface receptors that was more common in locally advanced tumors. The mode of clinical management (radiation versus chemotherapy) contributed distinct mutational signatures yet these mutations are not predicted to have functional relevance to tumor progression. Overall, these findings provide a first definition of the genetic features that distinguish among patients with locally advanced versus metastatic PDAC. These findings may have clinical relevance in upfront clinical decision making for the optimal candidates for neoadjuvant therapy.

## Introduction

Pancreatic ductal adenocarcinoma (PDAC) is a complex disease with an aggressive clinical course^1^. The complexity of PDAC stems from multiple factors unique to this tumor type. First, PDAC arises through the accumulation of somatic alterations in potent and currently undruggable driver genes^2^. Second, genetic events are also linked to transcriptional subtypes, some of which abrogate the efficacy of standard of care management^3^. Third, the PDAC ecosystem contributes to aggressiveness through its combination of cancer associated fibroblasts that influence cancer cell behavior, the formation of abundant extracellular matrix that increases intratumoral pressure and reduces vascularization, and a highly immunosuppressed environment^4^. For these and other reasons most patients are diagnosed with advanced stage disease^5^. Advanced stage PDAC comprises patients diagnosed with locally advanced non-metastatic (LAPC, Stage III) or metastatic PDAC (mPDAC, Stage IV). Within these two groups there exists heterogeneity of responses to treatment, or of metastatic efficiency. For example, some patients with LAPC have tumors that are downstaged to resectable status following a course of neoadjuvant therapy^6^, whereas others with LAPC will develop metastatic failure in one or more organs within a year of completing chemoradiation. Among patients with mPDAC, the range of metastatic burden during clinical management can vary one log fold^7,8^.

PDAC, like all forms of cancer, is an evolutionary process^9^. One goal of evolutionary cancer biology is to unveil the principles underlying cancer progression which remains the proximate cause of cancer related death^10,11^. At the genetic level, clonal driver mutations are regarded as the basis of tumor development while subclonal drivers are responsible for disease progression and treatment resistance^11,12^. Evolutionary and population genetic theories also propose that pre-existing resistant subclones expand under the selective pressure imposed by treatment and subsequently replace the predominant clones^9,13,14^. However, solid tumors that undergo neutral evolution are an exception to this notion in that few if any subclonal drivers accumulate that contribute to disease course in a meaningful way^15^. Collectively, the success of personalized cancer medicine requires proper characterization of mutations in the context of varying clinical presentations and treatments. Such characterizations should be considered in a tumor-type specific manner as well given different tumor types have different strategies for clinical management^16^.

Several large-scale genomic studies of PDAC have been reported that have predominantly focused on resectable PDACs^17,18^. The goal of this study was to perform a similar type of analysis but for patients who are diagnosed at advanced stages of disease (Stage III and Stage IV); such patients represent the up to 85% of new diagnoses^1^. Unlike our prior study of treatment naïve Stage IV PDAC^19^, the patients of interest in this study all received chemotherapy or combined chemotherapy and radiation. Our aim is to reveal how the genomic landscapes differ with respect to the stage of diagnosis and as a result of their corresponding standard of care therapies, as well as amongst chemotherapeutic agents that have different modes of action. To this end, we characterized the somatic mutation spectrum and the mutational signatures driving PDAC initiation and progression using next-generation sequencing-based cancer genome analysis.

## Results

### Clinical Features of Advanced Stage Patient Cohort

We sequenced whole exomes of 240 distinct tissue samples obtained through multi-region sampling of the primary tumor (120 distinct samples), matched metastases (90 distinct samples) and one normal tissue from 30 patients who underwent a research autopsy (**Supplemental Table 1 and 2**). Twenty-three patients (77%) were female, and the average age was 66.2±14.2 years. Most patients (60%) were originally diagnosed with Stage IV (metastatic) PDAC. The remaining patients were all diagnosed with nonmetastatic disease, most commonly Stage III PDAC. Two patients were diagnosed with Stage II disease; one (PAM31) opted for nonsurgical management whereas the second (PAM18) was resected but declined adjuvant chemoradiation.

Patients were categorized into three groups based on their treatment history (**Figure 1a, b, Supplemental Table 1**). Our rationale for this grouping is that the clinical management of each patient may reflect nuances of each patients’ disease not fully captured by clinical stage alone, i.e. if locoregional control was of primary concern in a patient with low burden oligometastatic disease. The first group of 13 patients, PAM19-PAM31, received chemoradiation for locoregional control of their primary tumor. Radiotherapy consisted of 50.4 Gy of radiation in 28 fractions with exception of one patient (PAM30) who received stereotactic body radiation therapy (SBRT, 33 Gy over five fractions). The concurrent systemic treatment in this group varied but most often consisted of 5-flourouracil or gemcitabine/oxaliplatin. A second group of 10 patients (PAM47-PAM56) received first line chemotherapy for de novo mPDAC, most often Gemcitabine with or without other agents. Finally, for comparison we included seven patients who never received systemic treatment, most often due to rapid disease progression or poor performance status at diagnosis. This group included patient PAM18.

**Figure 1.**
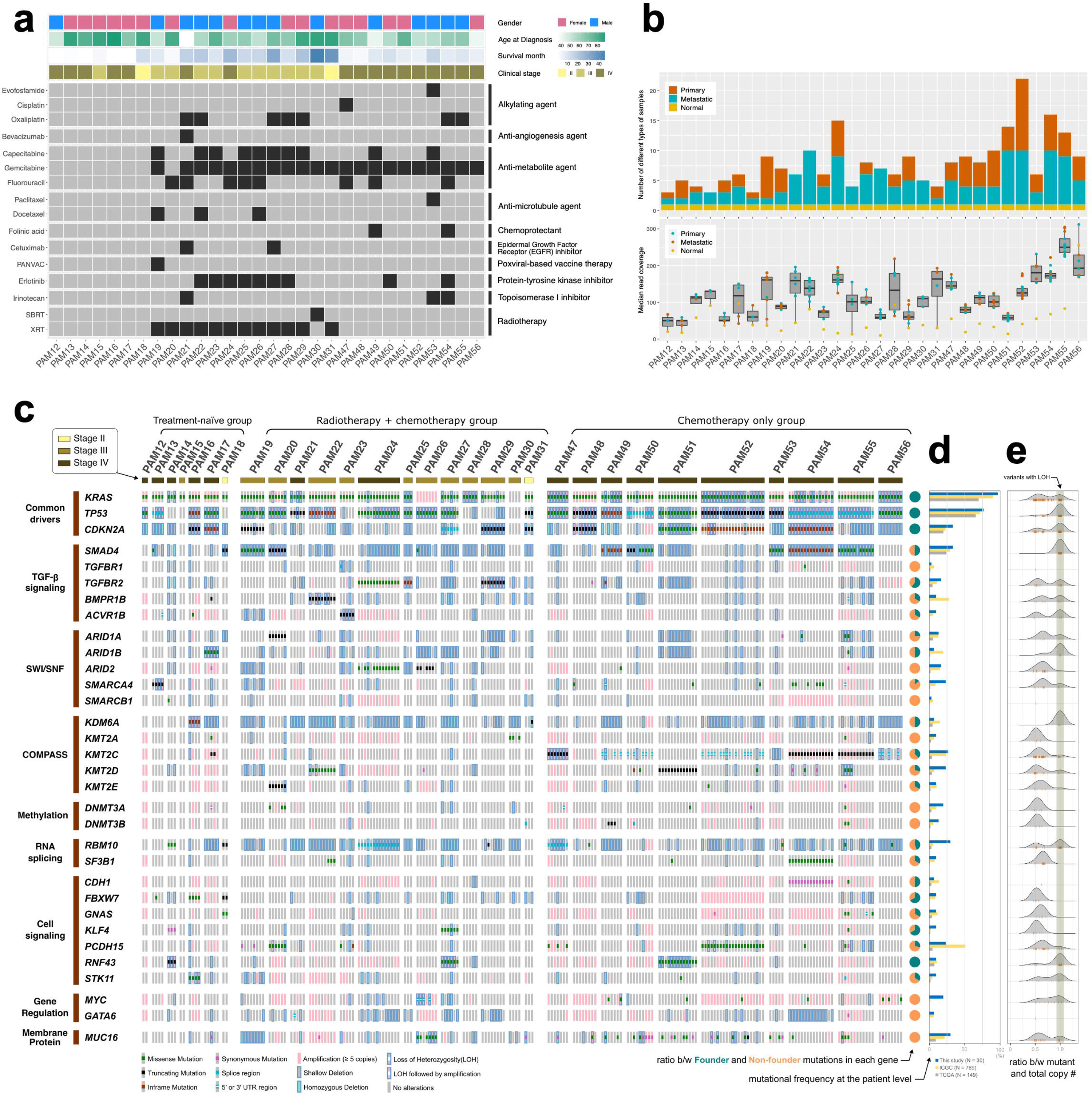
**a. Summary of clinical information and treatment plans for all PDAC patients in this study.** **b. Summary statistics of samples in each patient and read coverage for WES sequencing data.** **c. Oncoprint of all major driver genes in this cohort.** **d. Comparison of frequencies of altered driver genes between this study and public databases including ICGC and TCGA.** **e. Pattern of Loss of Heterozygosity (LOH) in each PDAC driver genes based on this study**

### Genomic Features of Advanced Stage PDACs

We identified 64,544 high quality somatic variants in total across all 210 cancer tissues (**Supplemental Table 3**). <u>Li</u>kely <u>F</u>unctional <u>D</u>river (LiFD) scores of all somatic mutations were then calculated to determine those most likely to be functionally deleterious (see **Methods and Supplemental Figure 1**). This filter resulted in 792 somatic mutations in 32 driver genes with an LiFD score of >0 in all samples, corresponding to 99 unique driver-like variants. An oncoprint of these driver genes reveals the expected high frequency of *KRAS* and *TP53* mutations followed by *CDKN2A* and *SMAD4* as the next two most commonly altered genes (**Figure 1c**). Recurrently altered genes that are seen at low frequencies in large cohorts were similarly identified that affected TGFβ signaling (i.e. *TGFBR1/2*), chromatin regulation via the SWI/SNF (i.e. *ARID1A, SMARCA4)* and COMPASS complexes *(i.e. KMT2C, KMT2D*), or RNA splicing (*SF3B1, RBM10*), among others. While the somatic alterations in this dataset are highly comparable to previously published findings in TCGA and ICGC, we note that for many genes the frequency is higher in this cohort (**Figure 1d**), likely due in part to more extensive sampling of each PDAC.

For each somatic mutation we next determined the extent that they were identified in all samples analyzed for each PDAC. We find that somatic alterations in only four genes, *KRAS, TP53, CDKN2A* and *RNF43*, are ubiquitously present in all samples of the PDACs they were found in. This pattern alone does not conclusively inform if mutations in these genes are founder mutations, i.e. present in the original cell that gave rise to the invasive PDAC. For this reason, we calculated the Cancer Cell Fraction (CCF) of each somatic mutation in all tumor samples for each patient. When categorizing mutation patterns in this cohort based on both multiregion sampling and CCF values per sample we find that *KRAS, TP53, CDKN2A* and *RNF43* are founder mutations in all of the PDACs in which these genes are mutated (see pie chart in **Figure 1c and Supplemental Figure 1**). By contrast, *SMAD4* is a founder mutation in only 50% of PDACs (5/10) in which it occurs. Nineteen additional driver genes that include *TGFBR2, ARID1A/B, KMT2C/D* and *FBXW7* were also found to be subclonal in a subset of the PDACs they were mutated in, whereas genes such as *ARID2* and *SMARCB2* are exclusively subclonal (see ‘Clonality’ column in the **Supplemental Table 3)**.

Copy number alterations (CNAs) affecting these 32 genes were common events across the cohort and exhibited frequent intertumoral and intratumoral heterogeneity (**Figure 1c,e**). For example, in addition to activating mutations we find frequent allelic imbalance and/or amplification of *KRAS*. In patient PAM26 amplification of wild type (WT) alleles was the sole mechanism identified for *KRAS* activation. More importantly, loss of heterozygosity (LOH) of the WT allele is highly prevalent in the select driver genes associated with PDAC evolution such as *TP53*, *CDKN2A* and *SMAD4* (**Figure 1e**). Homozygous deletions of these and other genes were also found. Haploinsufficiency was common for *KDM6A* and *RBM10*, irrespective of the presence of a coding somatic alteration, consistent with their location on the X chromosome. For genes such as *MYC* or *GATA6*, copy number alterations were the predominant mechanism of somatic alteration.

### Mutual Exclusivity of TGFβ Pathway alterations

*SMAD4* plays a critical role in canonical TGFβ signaling in epithelial tissues and is genetically inactivated in ∼50% of PDAC xenografts by mutations coupled with allelic loss or by homozygous deletion^20^. We sought to better understand the clonal and subclonal features of *SMAD4* inactivation in greater detail. *SMAD4* inactivating mutations were found in two of seven untreated PDACs (29%), three of 13 PDACs treated with chemoradiation (23%), and five of 10 PDACs treated with chemotherapy alone (50%) (see **Figure 1c**). These mutations corresponded to both clonal (founder) and subclonal events, and in all patients were accompanied by loss of the WT allele. The adjusted CCFs of *SMAD4* approximate 1 in most primary tumor samples and metastases in which it was found suggesting a relatively strong selective advantage for PDAC cells with these mutations (**Figure 2a**). Homozygous deletions were also noted in 6 patients, all of which were subclonal. In 10 PDACs both somatic mutations with LOH and homozygous deletions were coexistent indicating convergence for loss of *SMAD4* (see **Figure 1c, e**).

**Figure 2.**
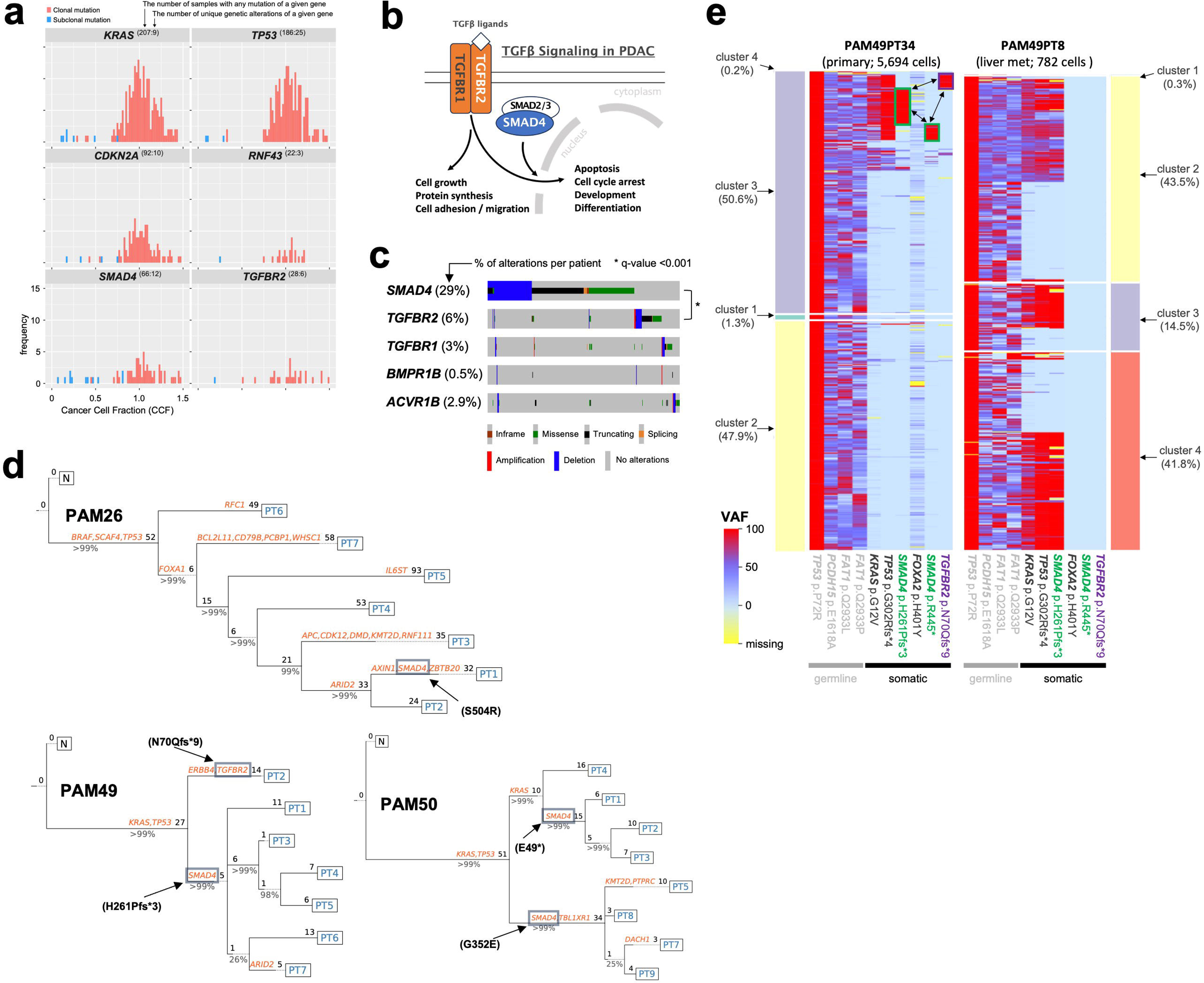
**a**. **Frequency distribution of estimated CCF values for four top PDAC driver genes** (see **Supplemental** Figure 2 for all major driver genes) **b**. **A brief overview of TGF**β **signaling pathway in PDAC**. **c**. Statistical testing of mutual exclusivity between *SMAD4* and *TGFBR2* mutations based on MSK-IMPACT targeted-seq data for selected 1,001 PDAC patients. d. Examples of phylogenetic tree of multi-region samples of selected patients (PAM26, PAM49 and PAM50) which have unique subclonal driver mutations in *SMAD4* and/or *TGFBR2*. e. Single cell genotyping maps of two samples from PAM49 as an example showing mutual exclusivity between *SMAD4* and *TGFBR2* in PDAC.

In light of this finding, we expanded our review of all mutations in the TGFβ signaling pathway^21,22^ (**Figure 2b**). Nine additional PDACs that were WT for *SMAD4* had mutations in the membrane receptors *TGFBR1, TGFBR2, ACVR1B* and *BMPR1B.* In five of these PDACs the mutations were founder mutations; all five of these PDACs were also primarily managed by chemoradiation; four patients were diagnosed with LAPC and one with oligometastatic mPDAC. Unlike *SMAD4*, mutations in these four surface receptors were not consistently accompanied by LOH (see **Figure 1e**) suggesting phenotypic consequences of haploinsufficiency alone as has been demonstrated in murine models^23,24^. Collectively, these findings indicate three previously unrecognized features of PDAC. First, convergence for loss of TGFβ signaling is a pervasive feature of PDAC; second, loss of signaling due to mutations in membrane receptors appears a feature of PDACs specifically with locally aggressive growth; and three, convergence for inactivation of TGFβ signaling occurs in both clonal and subclonal manner. Based upon reviewing available data for 1,001 PDAC patients that underwent targeted sequencing with MSK-IMPACT (see **Figure 2c**), we found that *SMAD4* and *TFGBR2* are mutually exclusive with statistical significance (q-value < 0.001 in a 2-sided Fisher exact test).

To understand the nature of subclonal driver gene mutations we generated sample phylogenies of patients in which this was observed (see **Figure 2d**). In patient PAM26 a single region of the primary tumor (Stage III at diagnosis) had a *SMAD4* mutation with CCF of 22% (see **Supplemental Table 3**). In PAM49, a *SMAD4* (p.H261Pfs*3) and a *TGFBR2* (p.N70Qfs*9) mutation are all non-founder ones and also located in different branches from the evolutionary tree. In PAM50 two *SMAD4* mutations were found (p.E49* and p.G352E) that were mutually exclusive from each other and present in clonal or subclonal CCFs amongst spatially distinct samples. This suggests that each mutation arose independently expanded and seeded different sites within the patient. Some sites shared both mutations suggesting polyclonal seeding.

We further validated the mutual exclusivity between *SMAD4* and *TGFBR2* mutations in PAM49 using single cell genotyping assay, Tapestri^®^ (Mission Bio). To this end, we chose one primary (PAM49PT34; 5,694 single cells) and one liver metastatic sample (PAM49PT8; 782 single cells) from the same patient for selected loci of known PDAC drivers based on our WES bulk-sequencing analysis (**Figure 2e**). First, the primary sample in PAM49 contained both *SMAD4* and *TGFBR2* mutations that are mutually exclusive as suggested based on the bulk WES data. Interestingly, this primary site contains different version of *SMAD4* mutation (p.R445*), which was not detected in the bulk sequencing data, being largely mutually exclusive with other *SMAD4* and *TGFBR2* mutations while colocalizing with *KRAS* and *TP53* (see the colored rectangular boxes and arrows in **Figure 2e**). On the other hand, the liver metastatic sample contained only one type of *SMAD4* (p.H261Pfs*3) but no *TGFBR2* mutations with very good colocalization with the other founder mutations such as *KRAS* (p.G12V) and *TP53* (p.G302Rfs*4), suggesting monoclonal seeding in the liver from the primary site.

### Treatment specific genetic alterations

The above data indicates that some genes are founders and pre-date exposures to cytotoxic or DNA damaging therapies used in PDAC management. However, the extent that these treatments themselves cause somatic alterations with deleterious effects is unknown.

We first compared the pattern of CCF estimates across different treatment plans in order to discriminate genetic alterations directly induced by each of treatments from the ones associated with PDAC initiation and progression (**Figure 3a**). The higher CCF a mutation possesses, the more likely it is to be a clonal mutation that occupies the majority of the cancer cell population^25^. We find that CCF estimates are positively correlated with LiFD scores (**Figure 3a and Supplemental Figure 2**) implying that a clonal mutation tends to have more functional impact than a subclonal one in PDAC. We find that the treated patients’ tumors tend to have more significantly increased proportion of subclonal mutations (less than 0.5 of CCF) than the naïve group while the clonal mutations having higher LiFD score are shared by all treatment groups (**Figure 3a**). Of note, a significantly larger number of subclonal mutations having LiFD > 0 exist in most treated patients unlike the non-treated ones (see the colored vertical bars on the x-axis in **Figure 3a**).

**Figure 3.**
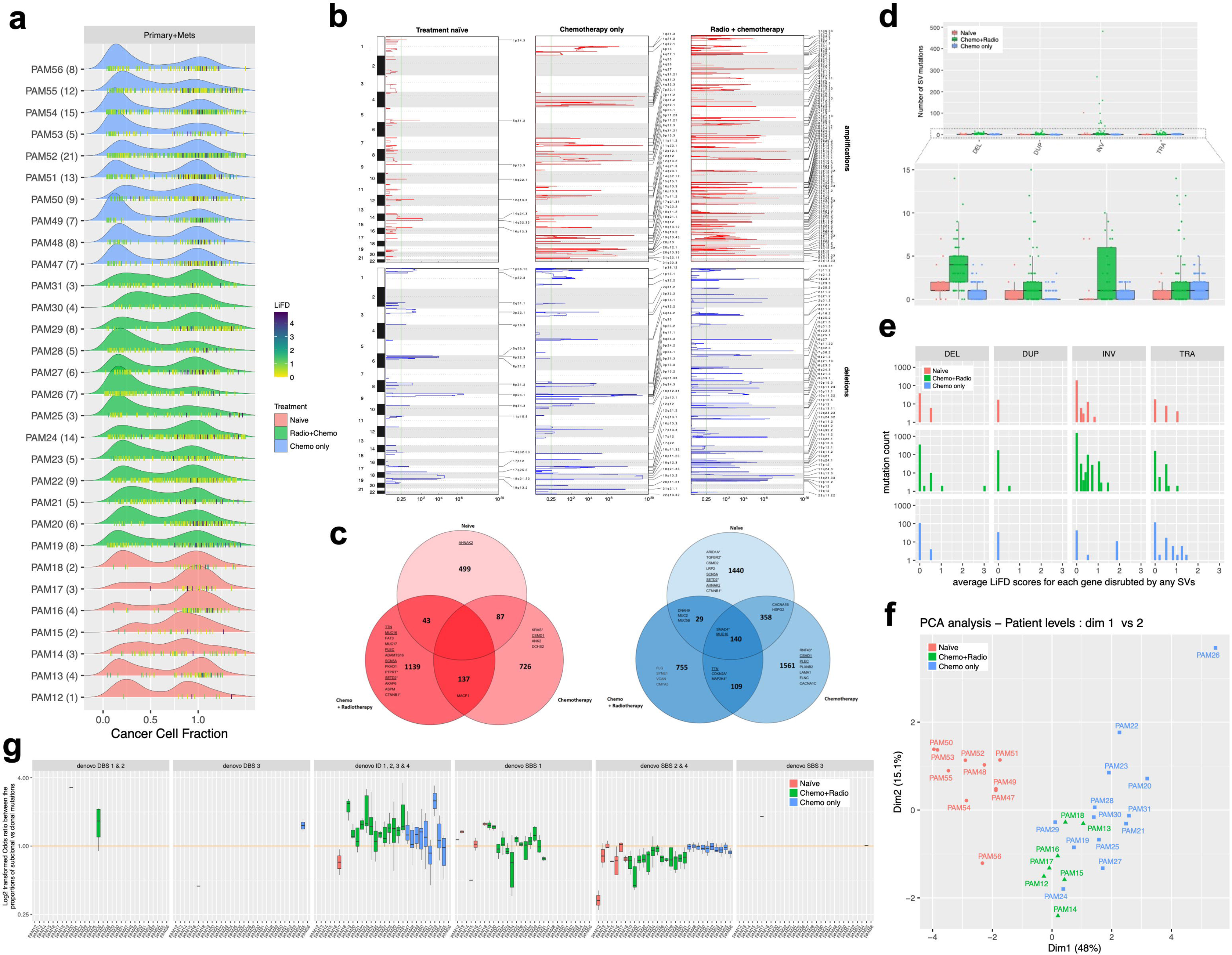
**a. The distribution of CCF estimates at the patient level**. Only samples with greater than 0.3 of purity were considered for reliable CCF estimates. The number in the parenthesis represents the number of samples in each patient included for this plotting. Bottom vertical bars represent the values of CCF and LiFD. **b. Original GISTIC2.0 results for each group**. Treatment naïve group includes 25 samples from 7 patients, chemotherapy only group includes 103 samples from 10 patients, and radiation plus chemotherapy group includes 83 samples from 12 patients. X- and y-axes represent q-value and chromosome, and the vertical green bar denotes the cutoff for significant peaks in gistic2 analysis. See **Supplemental** Figure 4 for selected genes of importance in PDAC, *MYC*, chromatin modifier and DNA repair pathways. **c. Comparison of the number and type of genes located in significantly amplified and deleted copy number regions in three treatment groups** (left panel: amplified genes, right panel: deleted genes). All the listed genes are ones that frequently acquire non-synonymous SNPs in PDACs based on cBioPortal (minimum number of sample occurrence is 20 for these genes). Genes with (*) denotes highly likely cancer driver genes additionally defined in OncoKB database. Some genes with underline mark(_) are amplified in some samples but deleted out in other samples within the same treatment group. **d. The number of different types of Structural Variants found in three treatment groups.** Each dot represents a single sample in each patient and treatment group. A box plot is drawn for entire samples in each group. (DEL : large deletions, DUP : large duplications, INV : inversions, TRA: translocations). Mutation count in the Y- axis is limited to 15 for simplicity. **e. Distribution of LiFD scores for selected genes that are disrupted by the breakpoints of any SVs.** **f. PCA analysis for mutational signatures at the patient level for three different treatment plans.** The average numbers of mutations corresponding to each signature was used for patient level analysis. Only mutations within exon or at 3’ / 5’ UTR are included. See **Supplemental** Figure 5 for PCA analysis result at the level of all samples including FFPE kept ones (PAM17PT3/PT4) and sequencing artifact ones (PAM55PT3/PT4) **g. Odds ratio between the proportions of subclonal versus clonal mutations.** We excluded samples that acquired less than 20 mutations both in founder and non-founder categories for each signature in this analysis in order to minimize over-representation of odds ratios.

Next, we identified the list of genes that are significantly amplified or deleted per treatment group by combining all tumor samples in each group and processing them together with the GISTIC2.0 (**Figure 3b** and **Supplemental Table 4**). We also highlighted driver-like genes that are found to frequently acquire non-synonymous SNPs in PDAC samples based on cBioPortal database (**Figure 3c**. see gene names in each Venn diagram). Overall, radiation clearly induced more frequent copy number gains and losses compared to PDACs that were not irradiated. The treatment naïve PDACs show almost no amplifications but significant number of deletions albeit to a lesser extent compared to the other groups. There are no single genes showing copy number gain commonly to the three patient cohorts. Many genes amplified only in chemo plus radio therapy group - *TTN*, *MUC16*, *PLECT*, *SCN5A*, and *SETD2* - are also deleted out in some samples in other groups. Given that most amplified genes only in radiation group are not driver-like, we suggest that radiation might increase the chance of occurrence of random copy number gain events across all chromosomes possibly with no or marginal phenotypic consequences. On contrary, many of genes including *SMAD4* were deleted out commonly in all three groups, representing that loss of function in *SMAD4* is an early driver rather than being induced by therapy.

Structural variants (SVs) are another type of mutational signatures contributing to tumorigenesis^26,27^. Using delly package^28^ with our bulk sequencing data, we defined four classes of somatic SVs: (1) large deletions (20+bp), (2) large duplications (20+bp), (3) balanced inversions and (4) translocations. First, we find that radiation treated group shows relatively large number of deletions and inversions (**Figure 3d**). Radiation-induced inversions are known to be frequently enriched in second malignancies regardless of cancer types^29^. We also confirm that inversions are the primary type of SVs induced by radiation compared to other types of SVs unlike other treatment regimens from our dataset.

We then investigated whether each type of SVs potentially link to cancer-driverness by looking up LiFD scores of each gene disrupted at its exonic regions by any of breakpoints of each SVs. First, each of different SVs didn’t show any noticeably different distribution of LiFD scores across different treatment groups (**Figure 3e**). In addition, majority of inversions found in the radiation group tend to have no sign of functional consequences in well-known PDAC driver genes based on LiFD scores, ranging mostly from 0 to 1. Of note, two radiation group patients - PAM20 and PAM26 - have massive number of inversions (up to 500) unlike the others in the same group which have around 10 inversions on average (**Supplemental Figure 3**). These patients also obtained much larger number of all types of somatic SVs possibly due to impairment in DNA repair mechanisms. It is unclear whether this hyper-mutated phenotype arose before or after radiation, but majority of these SVs might have no cancer-driving effects given their very low LiFD scores (**Figure 3e**).

Mutational signature analysis is an effective way of identifying unique mutational processes associated with different treatment strategies^30^. To this end, we evaluated three different types of signatures: 1) Single Base Substitution (SBS), 2) Double Base Substitution (DBS) and 3) Insertion and Deletion (ID) based on COSMIC Mutational signatures version 3^31^ using the R package ‘Palimpsest’^32^. There are eleven *de novo* signatures found in total: four *de novo* SBS, three *de novo* DBS and four *de novo* ID signatures (see **Supplemental Figure 4**). PCA analysis was performed at the sample and the patient level with the eleven signatures in order to correlate each unique treatment regime with the number of *de novo* signatures for each sample and patient (**Figure 3f and Supplemental Figure 5**). We find that radiotherapy treated samples differ from non-radiotherapy treated ones in terms of the number of total mutations for each of eleven *de novo* signatures, suggesting that radiation induces significantly increased mutational process within tumor tissues.

From an evolutionary perspective, founder (or truncal) mutations in the phylogenetic tree are likely an early driver initiating tumorigenesis^33^ and thereby shared by most tumors across different patients while non-founder ones are possibly occurring as responses to a strong selective pressure induced by treatments. There is, however, limited knowledge about the differences in acquisition of non-founder mutations depending on the type of treatment. To study this, we first classified all the somatic mutations into founder versus non-founder ones (the latter including both private and branched mutations) based on the phylogenetic tree topology for each individual patient. We reclassify eleven *de novo* signatures into five representative ones based on the cosign similarity of each signatures (**Supplemental Figure 6)**, and then assigned the highest ranked *de novo* mutational signature to each variant. Finally, odds ratio between the number of non-founder versus founder somatic variants in each representative *de novo* signature shows the existence of distinct adaptive evolution trajectory. We find *de novo* ID signature 1, 2, 3 & 4 was significantly over-represented in non-founder mutations compared to founder ones mainly in patients group treated with radiation (see green colored box plots in **Figure 3g**). Theses *de novo* ID signatures highly correlates with COSMIC ID8, a DNA repair of double strand breaks induced by radiation (see **Supplemental Figure 5**). We couldn’t find any other significantly altered mutational signatures in between founder vs non-founder classes under any other treatment regimes.

### Phylogenetic and pathway analysis of PDAC tumors in different treatment groups

We performed a sample level phylogenetic analysis of the multi-region samples sequenced for each patient with Treeomics^34^. In general, majority of driver mutations such as *KRAS*, *TP53* and/or *CDKN2A* are located in the trunk of each evolutionary tree in all treatment groups, indicating that they are common founders in PDAC progression (**Figure 4a**). The topology of each tree is, however, differs between patient groups who did or did not receive any therapy (**Figure 4b**). Patient groups treated with both radiotherapy and chemotherapy show massively increased proportion of non-founder mutations - somatic variants uniquely found in each individual sample - along with highly elevated deviation in that number within different samples from the same patient compared to other groups. The similar pattern of increased proportions of non-founder variants is also seen in patients treated only with chemotherapy albeit to a much less extent while no dramatic deviation in that proportion across samples was observed. This implies that all types of treatments might induce more private mutations in general and that radiation elevates the rate of mutational occurrence in a random way at each tumor sites.

**Figure 4.**
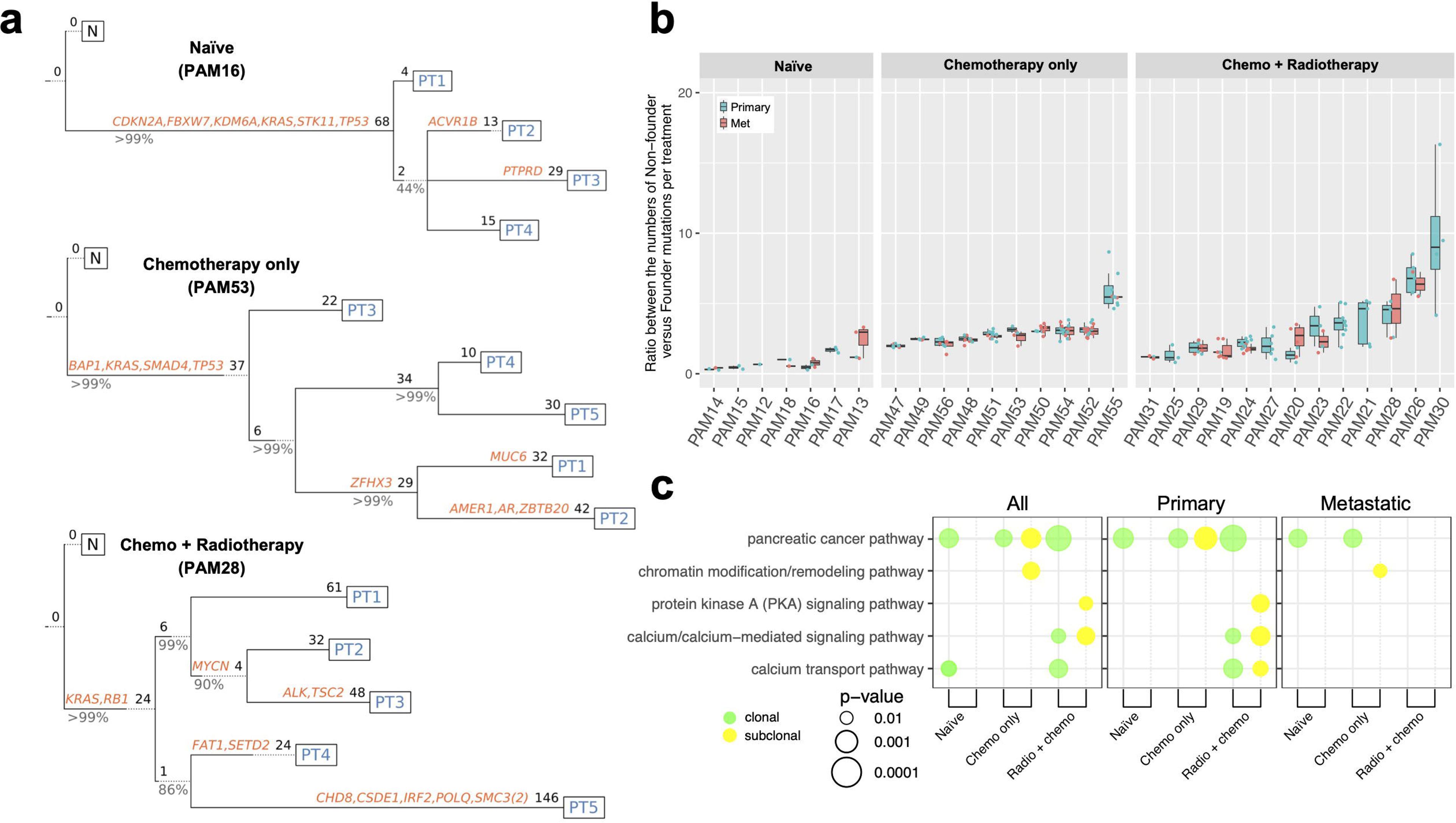
**a. Representative phylogenetic trees of each treatment plan.** **b. Ratios between the numbers of non-founder vs founder mutations in primary and metastatic samples per patient and treatment plan.** **c. Statistical significance of altered pathways in different treatment groups.**

We also figured out whether clinically defined biological pathways are associated with clonality and the treatment plans each patient experienced using RGD pathway database (https://rgd.mcw.edu/wg/home/pathway2/)^35^ (**Figure 4c**). First, we selected driver-like genes acquiring at least one somatic alteration(s) with greater than 0 of LiFD score in any of three different treatment groups and assigned them to each of 41 molecular pathways selected from RGD database, which are top 3 hierarchical ones having at least 200 genes (**Supplemental Table 5**). A two-sided fisher-exact test was applied to calculate p-values based on a contingency table containing numbers of the associated vs the non-associated genes for a given pathway of interest and all other exclusive pathways. We finally determined five RGB pathways are significantly altered by splitting the list of mutated driver genes in each treatment groups into clonal versus subclonal ones, which is defined by CCF estimate using Palimpsest in R: (1) pancreatic cancer pathway, (2) chromatin modification/remodeling pathway, (3) protein kinase A (PKA) signaling pathway, (4) calcium/calcium-mediated signaling pathway, and (5) calcium transport pathway (**Figure 4c**).

We find that all treated groups show any of significant subclonal pathway alterations (yellow circle in **Figure 4c**) whereas treatment naïve group didn’t. Such subclonal evolution in treated groups is less prevalent in metastatic samples compared to primary ones, consistent with the fact that metastasis in solid tumors develops through an evolutionary bottleneck induced by a stringent therapeutic selective pressure^36^. Our data demonstrates that pancreatic cancer pathways is significantly altered for clonal group of mutated genes such as *KRAS*, *TP53*, *SMAD4*, *CDKN2A* and others in all treatment groups as expected. Interestingly, the chemotherapy only treated group showed highly likely subclonal enrichment for those cancer-related pathways only for primary samples but not metastatic ones. However, no subclonal cancer-related pathways are shown in either cases where additional radiotherapy was applied, or no treatment was given (**see Figure 4c and Supplemental Table 5**).

## Discussion

While the recurrent genetic alterations of PDAC have been well described ^2,11,12^, the extent to which these genes occur in a subclonal manner or signify convergent evolution has not. These data now clarify the extent of convergent evolution of key signaling pathways in PDAC. For example, we find that virtually all genetic targets in PDAC occur in a subclonal manner in at least one PDAC; some genes such as *ARID2* were exclusively found in a subclonal manner whereas others were exclusively clonal (founders). Given the size of this cohort the extent that these frequencies can be extrapolated to early-stage disease are unknown.

A second major finding of this work is that the TGFβ signaling pathway, a known and common genetic target in PDAC, is subject to pervasive convergent evolution at both the intertumoral and intratumoral levels. *SMAD4* remains the most common genetic target and in this cohort its inactivation segregated with PDACs with high metastatic efficiency. However, our data further show that *SMAD4* WT PDACs are enriched for genetic inactivation of TGFβ surface receptors, most often *TGFBR2*, followed by *ACVR1B, BMPR1B* and *TGFBR1*. We also noted that mutations in TGFβ surface receptors were enriched within PDACs diagnosed at Stage III, i.e. locally advanced non-metastatic disease. This finding suggests that there is a genetic basis to the locally advanced phenotype, specifically the cellular level at which TGFβ signaling is disrupted. *SMAD4* inactivation results in loss of intracellular signaling from activated TGFβ receptors to the nucleus that is mediated by *SMAD2/3* in complex with *SMAD4*, whereas loss of TGFβ surface receptors leads to loss of activation of the pathway from autocrine or paracrine signals. In light of the finding that the greatest source of TGFβ ligand is within the desmoplastic stroma, these findings suggest that stromal features serve as a selective pressure for PDACs cells with loss of TGFβ signaling.

The Cancer Cell Fraction (CCF) of somatic mutations represents the order of mutational occurrence: the higher CCFs a mutation possess, the earlier it might have occurred during cancer evolution^37,38^. Our analysis shows that CCFs of many well-known PDAC drivers in metastatic samples are highly comparable to the ones of primary sites from the sample patient regardless of types of treatments, suggesting that metastatic seeding rapidly occurred before treatments (early seeding)^36^. PAM50 patient in this study is an example of revealing the clonal seeding patterns or the timing of metastasis using CCF estimates with bulk sequencing data. There are two different *SMAD4* mutations (p.E49* and p.G352E) with varying CCF values in different organs in this patient (see **Supplemental Figure 7 and Supplemental Table 3**). First, one primary (PT4) has no *SMAD4* mutations. On the other hand, p.E49* mutation is clonal in two primaries (PT2 and PT3) with 100% of CCFs while p.G352E is subclonal in the same samples only with 15-20% of CCFs. This indicates that primary clones in this patient are a mixture of different competing *SMAD4* alleles - WT, p.E49* and p.G352E. More interestingly, p.E49* is found only in lymph node met (PT1) but not in other metastatic sites including peritoneum, large bowel, and omentum while p.G352E is found only in such abdominal sites at ∼100% of CCFs. Presumably, a minor clone bearing p.G352E in the primary metastasized only to the organs in the abdominal cavity while p.E49* clone metastasized only to the lymph node (an example of monoclonal seeding). Metastatic seeding to the lymph node might have occurred earlier than seeding to the abdominal cavity given that p.E49* is a major allele in the primaries and therefore a possibly an earlier driver during the course of tumor development.

The distribution of CCF estimates might also represent the evolutionary trajectories shaped by different selective pressures. We find that the clonal mutations having higher LiFD score are common regardless of the type of treatments each patient experiences. However, the treated patients’ tumors tend to have more significantly increased proportion of subclonal mutations (less than 0.5 of CCF) than the naïve group (**Figure 2c**). We find a significantly large number of subclonal mutations having LiFD > 0 exist in most treated patients compared to the non-treated ones (see the colored vertical bars in **Figure 2c**). Such a subclonal variant could potentially be a resistance mutation conferring selective advantage against each treatment rather than a random genetic alteration directly caused by each drugs or radiations.

By calculating CCFs with combination of bulk and targeted deep-sequencing data, we identified several cases where subclonal *KRAS* mutations co-exist with a clonal allele at the range of 10-90% of CCFs (See **Supplemental Table 3**). This was found from all type of treatment groups: PAM17 (a treatment naïve group), PAM22 (a radio plus chemotherapy group), PAM50 and PAM53 (a chemotherapy only group). All of these secondary *KRAS* alleles are not only subclonal in each sample but also non-founders given that they are found only in a subset of selected tumor samples for the same patient. This demonstrates that the early drivers like *KRAS* are competing even at the terminal end of tumor evolution and that it is critical to determine the relative fitness of various *KRAS* alleles in different microenvironment in the future study.

Other subclonal mutations found in this study are possibly associated with altered functions in various metabolic and signaling pathways for PDAC patients. Among such subclonal variants there could be a resistance mutation conferring selective advantage against each treatment rather than a random genetic alterations. For example, chromatin modification/remodeling pathway is well known for its impacts on tumorigenesis across a broad range of tumor types and has been considered a potential therapeutic target^39–41^. Interestingly, our data shows that this pathway is significantly altered only in metastatic samples treated with chemotherapy (see **Figure 4c** and **Supplemental Table 5**) and that most of related alterations are subclonal as reported in a previous pan-cancer genome-based gene set analysis^42^. This possibly indicates that the subclonal alterations in chromatin modification complexes occur after chemotherapeutic treatment and result in intra-tumor heterogeneity (ITH) and/or drug resistance.

## Methods

### Ethics statment

Use of all human samples in this study was approved by the Institutional Review Board(IRB) at Memorial Sloan Kettering Cancer Center (under protocols #15-149, #15-021) and Johns Hopkins Medicine. All the case IDs used in this study (PAM12-PAM56) does not reveal the identity of each study subject.

### Patient selection and tissue processing

A cohort of 30 patients from the Gastrointestinal Cancer Rapid Medical Donation Program at the Johns Hopkins Hospital was used for this study. Tumor samples were cut from frozen sections and reviewed from microdissection and H&E staining in order to select ones with high cellularity (at least ∼ 20%). DNA extraction was done using DNeasy Blood & Tissue Kits for frozen samples following the manufacturer’s guideline.

### Whole Exome Sequencing and custom targeted re-sequencing

Extracted DNAs were processed on an Illumina HiSeq 2500 in a 100×100 paired end mode using the Agilent SureSelect V4 chemistry in the Integrated Genomics Operation (IGO) at Memorial Sloan Kettering Cancer Center (New York, NY). Our target read coverage for tumor and matched normal samples were ∼300X and ∼100X, respectively. Raw FASTQ files were aligned to ‘hg19’ human reference genome using bwa 0.7.17^43^ to generate BAM files for each sample. Picard Tools v2.26.0 was used to post-process the BAM files by removing soft-clipping reads, tagging duplicated reads, and sorting all reads by coordinate (http://broadinstitute.github.io/picard/). Then, Mutect2 in GATK2 package^44^ was used to identify somatic mutations by comparing each tumor sample with a matched normal sample in each patient. All options in Mutect2 were set as default except that ‘normal-lod’ was lowered to 1 instead of 2.2 in order to minimize false negative calls due to shallow read coverage in normal samples. We selected mutations that have at least one read per each strand in paired end sequencing and that are located within 500 bps from each exon. To validate the mutations found from WES, the same samples were additionally sequenced in much deeper coverage (500x-1000x) using a custom targeted panel based on the list of all unique somatic mutations from the original WES and an Illumina HiSeq 2500 in a 100×100 paired end mode, and then processed via the same bioinformatics pipeline as WES. The allele frequencies of somatic mutations from targeted re-sequencing data were multiplied to the read coverage number from WES data in order to get adjusted mutant allele counts and rescue somatic mutations that were missed in WES due to their low read coverage. Ensembl Variant Effector Predictor (VEP) v105^45^ and MSKCC’s Variant-

PostProcess pipeline v3 (https://github.com/soccin/Variant-PostProcess) were used to determine the effect of the finally selected somatic mutations. We then additionally recued mutations that are filtered out by any of the filtering steps only in subset of samples in each patient since they might be a false negative simply due to their low read coverage (generally less than 10). Raw sequencing data for both WES and targeted re-sequencing have been deposited as a BAM file format in the European Genome-Phenome Archive (EGA) under accession number EGAS00001007379.

### Cancer Cell Fraction (CCF) estimation and mutational signature analysis

First, allele specific copy number of each somatic mutation was determined by FACETS^46^. Then sample purity of each tumor sample was calculated using FACETS (copy number based) and Ccube^47^ (snp-based), individually and then averaged as one. We filtered out somatic mutations with low quality by setting up the minimum coverage as 0.1X of the median read coverage of all mutations in an individual sample, the minimum tumor allele count as 2 and the minimum variant allele frequency as 0.5%. Palimpsest v2.0 ^32^ in R package was used to estimate CCF of each somatic mutation in an individual sample by feeding in the inferred allele specific copy number and the sample purity for the selected list of somatic mutations. Then, clonality of each mutation in each sample was calculated using default settings in the package. We also inferred *de novo* Single Base Substitution(SBS), Double Base Substitution(DBS) and Indel(ID) level mutational signatures for each variant at individual sample level using the same Palimpsest package. Formalin-Fixed Paraffin-Embedded (FFPE) tumor samples (such as PAM17T3,4) were excluded as a technical artifact since it induces distinct signature patterns as a result of DNA crosslinking. Specifically, ‘brunet’ method, one of Nonnegative Matrix Factorization (NMF) algorithm, was used to infer SBS and ID level signatures while ‘nsNMF (non-smooth NMF)’ algorithm was used for DBS level signatures estimation. The number of runs for each algorithm was set to 50. Final VAFs, clonality and *de novo* signature information of all somatic mutations are listed in the **Supplemental Table 3**.

### Cancer driverness estimation

Likely functional driverness specific to PDAC was predicted using LiFD package^48,49^, which is a two-phase algorithm that integrates multiple information from public cancer databases such as OncoKB, CGI and COSMIC and bioinformatic methods including CHASMplus, FATHMM, CanDrA+, CGI and VEP (https://github.com/johannesreiter/LiFD). Input file was ‘VCF’ formatted list of mutations and all settings used in the package were default. ‘LiFD_support’ score ranging 0 to 5 was selected as the final determinant of driverness of each mutation. Less than 1 of LiFD_support score is regarded as non-driver gene while higher score represents a more likely driver.

### Analysis of significantly perturbed copy number regions specific to each treatment plans

GISTIC2.0^50^ was used to infer the significantly amplified or deleted arm-level and focal copy number regions for each treatment group. First, we used FACETS to calculate chromosomal segment information of all somatic copy number variants by comparing each tumor’s WES data and its matched normal. The minimum and the maximum depth of normal sample were set as 35 and 4000, respectively. The minimum and the maximum critical value used for segmentation were set 25 and 150, respectively. All individual sample level segmentation files were combined for each treatment group and fed into GISTIC2.0 package as an input. The reference genome used was hg19, which is the same as the one used for WES data analysis. All other settings in GISTIC2.0 were default, and the cutoff of q-value was 0.25 to determine significantly perturbed copy number regions. We then extracted the list of all genes included in the significant copy number altered regions that are specific to each treatment plan.

### Analysis of Structural Variants (SVs) specific to each treatment group

We inferred individual tumor sample level SVs including large deletions / duplications, balanced inversions and translocations using ‘sv-callers’ workflow (https://github.com/GooglingTheCancerGenome/sv-callers), which utilizes multiple SV inferring packages such as Manta, Delly, Lumpy and GRIDSS simultaneously. Matched normal mode was selected and all other settings were default. We included only results from Delly^28^ since it outputs SVs with ‘PASS’ tag for our WES dataset while others didn’t. The minimum read coverage was set to 30 for final filtering. We further investigated which genes are disrupted by any given SV within exon and intron of the gene by looking up the SV’s coordinate.

### Analysis of significantly perturbed pathways specific to each treatment group

First, 142 human pathway annotation terms having at least one annotated gene under six top2-level pathways – classic metabolic pathway(PW:0000002), disease pathway(PW:0000013), drug pathway(PW:0000754), regulatory pathway(PW:0000004), and signaling pathway(PW:0000003) – are collected from RGD database^35^ (https://rgd.mcw.edu/wg/home/pathway2) (see **Supplemental Table 4**). We then split the list of somatic mutations for each treatment group into two classes: clonal vs subclonal one based on the column ‘clonality’ in the **Supplemental Table 3**, filtered out pathways having less than 200 genes in it and then separately applied Fischer Exact test based on a 2×2 contingency table as shown below to calculate statistical significance. The null hypothesis is that a chosen pathway is NOT different from other pathways for the number of genes in it for each given class (clonal or subclonal).

2 x 2 contingency table
|  | NOT included | Included |  |
| --- | --- | --- | --- |
| pathway A | **a** | **c** | a + c |
| NOT pathway A | **b** | **d** | b + d |
|  | a + b | c + d |  |

**a** : the number of genes that are present in this pathway 'A' & NOT included in a given class (clonal or subclonal)
**c** : the number of genes that are present in this pathway 'A' & included in a given class (clonal or subclonal)
**b** : the number of genes that are NOT present in this pathway 'A' & NOT included in a given class (clonal or subclonal)
**d** : the number of genes that are NOT present in this pathway 'A' and included in a given class (clonal or subclonal)

### Phylogenetic analysis of multi-region sampled tumors in each patient

Treeomics package^34^ was run to reconstruct the tumor patient’s phylogeny for the cases where the minimum number of tumor samples is 3 after excluding samples with less than 10% of computationally estimated tumor purity. For this reason, PAM12 and PAM15 were excluded for this analysis. Since the maximum number of tumor samples for this analysis is limited due to its high resource intensity, we selected only up to 12 tumor samples for each patient that are as much unique as possible compared to other tumors in the same patient by looking up Pearson correlation coefficient (PCC) of allele frequencies of all somatic mutations in each pair of tumor samples. The number of best solutions explored by ILP solver to assess the support of the inferred branches was set to 5,000 (default 1,000) and the maximum variant allele frequency for an absent variant before considering the estimated purity was set to 1% (default 5%). We only selected the best scored tree for each patient and visualized the total number of all mutations and the driver-like gene names according to the list of TCGA consensus driver genes^51^ at each branch line in black and orange color, respectively.

## Data Availability

All data produced in the present study are available upon reasonable request to the authors

